# Choosing questions before methods in dementia research with competing events and causal goals

**DOI:** 10.1101/2021.06.01.21258142

**Authors:** L. Paloma Rojas-Saunero, Jessica G. Young, Vanessa Didelez, M. Arfan Ikram, Sonja A. Swanson

## Abstract

Several of the hypothesized or studied exposures that may affect dementia risk are known to increase the risk of death. This may explain counterintuitive results, where exposures that are known to be harmful for mortality risk sometimes seem protective for the risk of dementia. Authors have attempted to explain these counterintuitive results as biased, but the bias associated with a particular analytic method cannot be defined or assessed if the causal question is not explicitly specified. Indeed, we can consider several causal questions when competing events like death, which cannot be prevented by design, are present. Current dementia research guidelines have not explicitly considered what constitutes a meaningful causal question in this setting or, more generally, how this choice justifies and should drive particular analytic decisions. To contextualize current practices, we first perform a systematic review of the conduct and interpretation of longitudinal studies focused on dementia outcomes where death is a competing event. We then describe and demonstrate how to address different causal questions (referred here as “the total effect” and “the controlled direct effect”) with traditional analytic approaches under explicit assumptions. Our application focuses on smoking cessation in late-midlife. To illustrate core concepts, we discuss this example both in terms of a hypothetical randomized trial and with an emulation of such a trial using observational data from the Rotterdam Study.

## 1. INTRODUCTION

Much research on dementia etiology focuses on understanding the role of biological factors in the pathophysiological process, and the impact of modifiable factors that could prevent or delay the onset of the disease[1]. As such, the field is often interested in causal questions. However, proper causal inference in dementia research faces many methodological and substantive challenges[2]. One of these challenges, which arises even in randomized trials, is that individuals at risk of dementia may die of other causes prior to its onset. In this setting, death is a *competing event* because an individual who dies from another cause prior to dementia onset cannot subsequently experience dementia[3].

Several of the hypothesized or studied exposures that may affect dementia risk can also increase the risk of death. This may explain counterintuitive results, where exposures that are known to be harmful for mortality risk, such as smoking[4,5] or history of cancer[6], sometimes seem protective for the risk of dementia. Authors have attempted to make sense of these counterintuitive results by naming biases such as “competing risk bias” or “survival bias”[7,8]. However, the bias associated with a particular analytic method cannot be defined or assessed if the causal question is not explicitly specified. We can envisage several causal questions when competing events like death are present, as we will discuss below.

Current dementia research guidelines[2] have not explicitly considered what constitutes a meaningful causal question in this setting or, more generally, how this choice justifies and should drive particular analytic decisions. Previous recommendations in the statistical and epidemiologic methods literature have advocated for the *cause-specific hazard* ratio when the aim is “etiologic”[9–12], relying on the popular Cox proportional hazards (PH) model. However, such recommendations cannot be justified (or criticized) without reference to the causal question which the analysis seeks to answer. The lack of explicit consideration of questions has contributed to confusion about the interpretation of different approaches to data analysis, including the misconception that “*censoring”* competing events is equivalent to somehow “*ignoring”* them. Recently, Young and colleagues placed core statistical concepts like *risk* and *hazard* within a formal causal inference framework and mapped common analytic strategies for competing events to questions about an intervention’s effect either with and without elimination of the competing event[13]. This work clarifies the role of censoring with respect to question formulation, assumptions needed for causal interpretation given real-world data, and particular analytic choices.

The goals of the current study are two-fold. To contextualize current practices, we first perform a systematic review of the conduct and interpretation of longitudinal studies focused on dementia outcomes where death is a competing event. Second, to contextualize the ideas formalized by Young and colleagues[13] in the setting of dementia research, we describe and demonstrate how to translate different causal questions (what we will refer to as questions concerning “the total effect” and “the controlled direct effect”) to popular analytic approaches under explicit assumptions. Our application focuses on smoking cessation in late-midlife, one of the twelve modifiable risk factors for dementia prevention described in the 2020 report of the Lancet Commission[1]. To illustrate core concepts, we discuss this example both in terms of a hypothetical randomized trial and with an emulation of such a trial using observational data from the Rotterdam Study[14].

## 2. SYSTEMATIC REVIEW OF LONGITUDINAL STUDIES OF DEMENTIA

### 2.1. Methods

We conducted a systematic review of recent original research articles with dementia outcome. We aimed to describe how death during follow-up is handled in the design, analysis, reporting, and interpretation.

Eligibility for our systematic review included: original research with longitudinal data on dementia or Alzheimer’s disease outcomes, published between January 2018 to December 2019. We limited our systematic review to nine journals in applied dementia or applied general medical research: Alzheimer’s and Dementia, Annals of Neurology, BMJ, JAMA, JAMA Neurology, Lancet, Lancet Neurology, Neurology, New England Journal of Medicine. We searched PubMed for papers that contained the words in the abstract: Alzheimer’s disease or dementia; longitudinal or cohort; hazard or risks (**Supplemental Data 1**).

We collected the following information from each eligible article: (1) reported study characteristics (type of exposure, median length of follow-up, study aim); (2) reporting on death and loss to follow-up (number of people who died over time, number of people who died over time by level of an exposure of interest, number of people lost to follow-up); (3) information on specific methodologic considerations (explicitly mentions how the competing event of death is handled in the analysis plan, primary target parameter, primary statistical method, explicitly mentions the assumptions needed for valid inference given the competing event of death and additional analyses and measures reported); and (4) interpretation (valid interpretation of the primary result given the competing event of death, discusses mortality in discussion).

### 2.2. Results

We retrieved 210 papers using the specified terms, 78 of which met eligibility criteria (**Supplemental Data 1**). Though we intended to classify articles according to the type of aim (descriptive, predictive and causal), this was not possible since most articles are not specific, and more frequently the term “association” is used to represent an ambiguous aim. Over 80% of the studies (n=63) reported associations between dementia and a single measure of a time-fixed or time-varying exposure (**Table 1**). Mean or median follow-up was over 5 years for 70% (n=55) of the studies. The number or proportion of individuals who died over time was reported in 41 (53%) papers; 12 (15%) presented these numbers by exposure level and 41 (53%) reported losses to follow up. Only 27% (n=21) explicitly reported how the competing event of death was handled in the methods section. The vast majority presented estimates of a hazard ratio (88%) based on a Cox proportional hazard model (85%). A table with the additional reported summary measures is present in **Supplemental Data 1**. Of all papers, only four explicitly mentioned assumptions pertaining to the presence of deaths by other causes (i.e., competing events). No papers that reported estimated coefficients of a (cause-specific) Cox proportional hazard model considered their explicit interpretation (see below)[3,13,15–18]. Overall, only one-third of the publications (n=25) mentioned death in some context (e.g., interpretation, limitation) in the discussion section.

**Table 1.** Current reporting practices relevant to competing events in dementia research (N=78 articles)

|  | N (%) |
| --- | --- |
| Exposure type |  |
| Not applicable | 2 (3%) |
| Time-fixed or time-varying measured at one time point | 63 (81%) |
| Time-varying | 4 (5%) |
| Time-varying treated as time-fixed | 9 (11%) |
| Median length of follow-up |  |
| 1 to 3 years | 7 (9%) |
| 3 to 5 years | 16 (21%) |
| 5 to 10 years | 24 (31%) |
| 10 to 15 years | 9 (12%) |
| 15 to 20 years | 5 (6%) |
| Above 20 years | 17 (22%) |
| Includes n or % of deaths | 41 (53%) |
| Includes n or % of loss to follow-up | 41 (53%) |
| Includes n or % of mortality By exposure level |  |
| No | 62 (79%) |
| Yes | 12 (15%) |
| Not applicable | 4 (5%) |
| Explicitly mentions how the competing event of death is handled in the analysis plan | 21 (27%) |
| Explicitly mentions the assumptions needed for valid estimation given the competing event of death | 4 (5%) |
| Primary statistical method |  |
| Cox-proportional hazard model | 66 (85%) |
| Cumulative incidence function | 3 (4%) |
| Fine-gray sub distribution hazard model | 2 (3%) |
| Multistate model | 1 (1%) |
| Poisson model | 3 (4%) |
| Other | 3 (4%) |
| Primary target parameter |  |
| Hazard Ratios | 69 (88%) |
| Rates (cases per person-year) – rate differences | 3 (4%) |
| Risk Ratios | 3 (4%) |
| C-statistic | 1 (1%) |
| Cumulative Risks (absolute risk - risk difference) | 1 (1%) |
| Sub-distribution hazard ratios | 1 (1%) |
| Explicit interpretation of the primary estimate given the competing event of death |  |
| No | 56 (72%) |
| No interpretation given | 18 (23%) |
| Yes | 4 (5%) |
| Mentions mortality in discussion section | 25 (32%) |

With the findings of this systematic review in mind, we next turn to describing important features of data structures with competing events, causal questions that can be posed, and under what assumptions and using what methods those questions can be answered.

## 3. FROM QUESTIONS TO METHODS IN DEMENTIA STUDIES WHERE SOME INDIVIDUALS DIE DURING STUDY: A PEDAGOGIC EXAMPLE

### 3.1. Observed data structure

Consider the effect of smoking cessation (versus continuing) in late-midlife on developing dementia after 20 years of follow-up. In order to focus on the challenges to causal inference created by competing events, let us begin by considering an idealized randomized trial such that middle-aged smokers are randomly assigned to a strategy to quit smoking versus continue smoking. Dementia onset is rigorously measured through constant screening, with date of death during follow-up recorded through linkage with municipal records. Further, suppose in the idealized trial we have complete follow-up (all individuals remain in the study until end of follow-up or until death) and perfect adherence.

Trial participants will be observed to follow different possible event trajectories through the study period: death without developing dementia; dementia onset (some dying after dementia onset); or remaining alive and dementia-free until end of follow-up. For those individuals who died without developing dementia, after the time of death, they cannot subsequently develop dementia. This is the key implication of competing events: they make it impossible for the event of interest to subsequently occur. It is precisely this determinism that makes choosing a causal question more difficult than when competing events are absent (e.g., when the event of interest is all-cause mortality rather than dementia).

The causal diagram in **Figure 1** represents some key features of this data structure, where an arrow from one node A into another node B on a causal diagram reflects that A may cause B[19]. Gaining familiarity with the key features in this causal diagram will illuminate the tradeoffs in interpretation of different definitions of a causal effect on dementia in the presence of death, as well as the assumptions used for identifying these effects from observable data. In this graph, *Smoking* represents an individual’s smoking status, and *Death*_*19*_ and *Dementia*_*20*_ represent indicators of death by 19 years of follow-up and dementia risk by 20 years of follow-up, respectively. By randomization, we know there are no shared causes of *Smoking* and other variables represented on the graph (the only cause of quitting smoking is a “coin flip”). However, we have no such guarantee for death and dementia status over the follow-up; therefore, the graph depicts shared causes *C* of dementia and death (such as cardiovascular comorbidities) that may or may not be measured. The arrows from *Smoking* to *Death*_*19*_ and *Smoking* to *Dementia*_*20*_ illustrate that smoking may affect both dementia and death through different mechanisms. The bold arrow from *Death*_*19*_ to *Dementia*_*20*_ represents the key feature of a competing events data structure: an individual who dies by year 19 of follow-up *cannot* subsequently develop dementia at the next time point, with the boldness here indicating the determinism. Though we present death and dementia at years 19 and 20 respectively, the causal diagram could be expanded to include their assessments in previous years as well. This simplified causal diagram is sufficient, however, for our consideration of the causal question.

**Figure 1.**
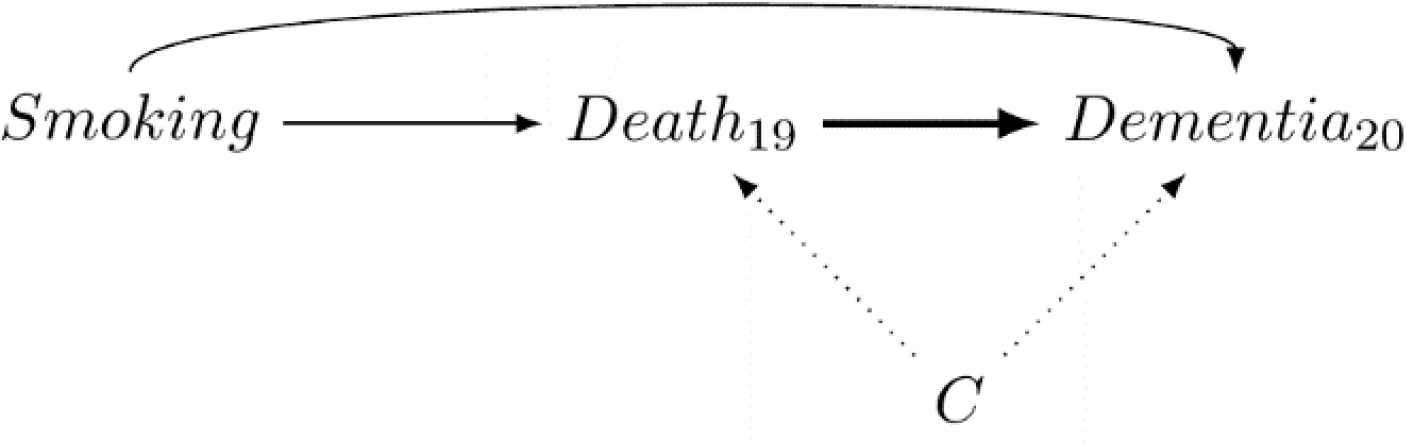
A causal directed acyclic graph representing some key causal features of the data structure. *Smoking* represents the exposure status (quit smoking vs. continue smoking), *Death(19)* and *Dementia(20)* represent indicators of death by 19 years of follow-up and dementia by 20 years of follow-up, respectively. *C* represents possible shared causes of dementia and death (such as cardiovascular comorbidities). The key relations are: 1) smoking may independently affect both the risk of dementia and death over time through different mechanisms; 2) dying over the first 19 years of follow-up (without prior onset of dementia) determines that the indicator of dementia at 20 years of follow-up is zero (the bold arrow representing this key determinism induced by competing events); and 3) dementia and death can have shared causes.

### 3.2. Choosing a causal question: the total and controlled direct effect

We say the study we have conceptualized is “ideal” because, in the case of a randomized trial with no loss to follow-up and perfect adherence, we can identify the exposure effect on the outcome of interest through all possible pathways: the *total effect*. In our example, the following is a question about a total effect: *What would the difference in dementia risk by 20-year follow-up be had all individuals in the study population quit smoking versus, instead, had all individuals continued smoking?* This dementia risk is an example of a “cause-specific cumulative incidence” or “crude risk”[3,17].

Unfortunately, the total effect captures all pathways by which exposure affects dementia, including those mediated by death. In the causal diagram in Figure 1 this includes both the *direct* effect on dementia (Smoking→Dementia_20_) and indirect of smoking via mortality (Smoking→Death_19_→Dementia_20_). This indirect effect is necessarily “protective” since participants who die due to smoking at an earlier time point are “protected” from developing dementia. This “pathological mediation” structure gives the total effect a potentially problematic interpretation, since smoking cessation may increase the risk of dementia but primarily or solely because it delays death. Thus, the total effect may not answer a desirable causal question in these settings, especially when there is an arrow between the exposure and competing event. Empirical support for this arrow and, in turn, for quantifying the concerns about “pathological mediation” of the total effect, can be obtained by also estimating the effect of smoking cessation on all-cause mortality.

Instead of a total effect, a direct effect of smoking on the risk of dementia (that does not also capture the pathways mediated by death) may be of interest. There are multiple ways to define a direct effect[20–22]. Here we will consider one definition that has been historically considered and may lead to familiar statistical methods as will be described in the next section: the *controlled direct effect*. In our example, this question is phrased as: *What would the difference in dementia risk by 20-year follow-up be had all individuals in the study population quit smoking <u>and not died throughout the study period</u> versus, instead, had all individuals continued smoking <u>and not died throughout the study period</u>*? This dementia risk (under elimination of death) is an example of a “net risk” or “marginal cumulative incidence”[3,17]. This effect only captures the direct effect of smoking on dementia because it refers to a hypothetical setting in which somehow death could be eliminated. As we discuss in the next section, while our idealized trial allows us to identify the total effect by design, it does not guarantee this for the controlled direct effect.

The risk differences above both quantify causal effects because they both refer to a comparison of outcome distributions under different interventions but in the same individuals. In contrast, while cause-specific hazard ratios are the basis of the majority of analyses in dementia studies, these generally do not quantify causal effects, even under the conditions of an ideal trial. Unlike risks, hazards are defined conditional on not yet having had the outcome or competing event. This conditioning means that hazard contrasts do not compare outcomes under different exposures in the same individuals when exposure affects these events[13,23,24]. Therefore, in dementia studies, cause-specific hazard ratios will not generally have a causal interpretation when exposure affects *either* dementia *or* death (directly or indirectly). For this reason, we will focus on risks even though many studies in our literature review reported hazard ratios.

In sum, there is no single way to define “the” causal effect on dementia when deaths occur. Choosing either of these research questions should be done in a case-by-case basis, and we will address this, as well as introduce other alternative questions, in the discussion section. Presenting information on the relation between exposure and mortality can complement both questions.

### 3.3. Identifying the total versus controlled direct effect in a real-world study

In this section we consider assumptions that help us connect our causal quantity of interest to observable data (i.e., identification). Consider again Figure 1: because exposure was randomized, there are no non-causal paths connecting *Smoking* and *Dementia*_*20*_[19,25]. This is consistent with the assumption of “no confounding”, allowing identification of the total effect. In contrast, to identify the *controlled direct effect* of smoking cessation on the risk of dementia, we need to make additional assumptions; that is, “no confounding” ensured by randomization of the exposure is not sufficient.

In Figure 1, we observe the non-causal path between death and dementia through their shared cause C, Dementia_20_←C→Death_19_. Thus, even in our ideal trial, we need to measure C to identify this effect. The reason we need to measure and adjust for C when interest is in the controlled direct effect is because death is a form of *censoring* for this question[13]. Censoring is a type of *missingness* in the outcome of interest. Therefore, what constitutes censoring depends on the question of interest. A, as such the controlled direct effect is a question about dementia outcomes in hypothetical settings where death is eliminated. When an individual dies prior to dementia onset, dementia onset “under elimination of death” is missing for that individual. While many researchers equate “death” with “censoring”, these terms are not synonymous: death is *only* a type of censoring (leading to missingness of the dementia outcome) when the question of interest is about outcomes “under elimination of death”.

In turn, measuring and including the shared cause C in Figure 1 of dementia and death is consistent with an assumption often referred to as *conditional independent censoring* (here, conditional on C) [3,10,11,13,17,18,26]. This assumption becomes more plausible in most studies if time-varying shared causes are included rather than only baseline covariates. Assuming that there are no shared causes between death and dementia (i.e., assuming the absence of the dotted arrows from C to Death_19_ and Dementia_20_ in Figure 1) coincides with the assumption of *unconditional independent censoring*. This is implausible for nearly all dementia research since both events are related to the aging process and consequences of it. Thus, even in our ideal trial, given interest in the controlled direct effect, we must measure a rich set of covariates related to both dementia and mortality risk at baseline and repeatedly throughout follow-up.

On a separate note, loss to follow-up is a form of censoring (missingness) whether interest is in total or direct effects. Though loss to follow-up can in principle be prevented by design, in trials or other studies of dementia, mechanisms of loss to follow-up might be related to impaired cognition and dementia[27], and as such, shared causes of loss to follow-up and dementia should be measured as well for similar reasons as for measuring C[28,29]. Further details on censoring and graphical identification of both effects, including scenarios with loss to follow-up, can be found in Young et al.[13].

### 3.4. Statistical methods to estimate the total effect or the controlled direct effect

Choosing an appropriate statistical method depends jointly on the choice of causal effect and the identifying assumptions we make. In an ideal trial, the *total effect* can be trivially estimated by simply comparing two proportions: the proportion diagnosed with dementia at 20-year follow-up in the “quit smoking” arm versus the proportion diagnosed with dementia at 20-year follow-up in the “do not quit smoking” arm. In both proportions, individuals who die before developing dementia will contribute to the denominator but never to the numerator. Likewise, these quantities can be estimated with the Aalen-Johansen estimator[13,17,18], which extends to settings with loss to follow-up under the assumption of unconditional independent censoring by loss to follow-up.

In contrast, the controlled direct effect requires covariate adjustment on the shared causes of death and dementia, even in an ideal trial. For example, the controlled direct effect could be estimated by comparing the risk estimates from the complement of a weighted version of the Kaplan-Meier estimator[13], where weights represent the inverse probability of censoring by death conditional on covariates[13,26,30–32]. These covariates should be those assumed to ensure the conditional independent censoring assumption for this form of censoring (e.g., the covariates C in Figure 1).

We note that the historic survival-analysis terminology classifies this structure as “semi-competing events” since death is a competing event for dementia but not the other way around. Therefore, we can estimate the risk of all-cause mortality using standard methods like the Kaplan-Meier estimator. In all cases, straightforward extensions exist for covariate adjustment (e.g., by inverse probability weighting) to address loss to follow-up as well as for confounding [13,29,33–35]. As such, these methods can be used in realistic trials and in observational studies, though our consideration of estimation in an ideal trial helps illuminate the unique feature of competing events.

## 4. APPLICATION TO THE ROTTERDAM STUDY

We now illustrate an application of inverse probability weighted methods to estimate total and controlled direct effects of smoking cessation on dementia using data collected from the Rotterdam Study. Briefly, the Rotterdam Study is a population-based prospective cohort study among persons living in the Ommoord district in Rotterdam, the Netherlands[14]. Participants older than 55 years underwent questionnaire administration, physical and clinical examinations, and blood sample collection at baseline (1990-1993) and at follow-up visits from 1993-1995, 1997-1999, 2002-2005, and 2009-2011. Smoking habits were assessed through questionnaires at study entry via self-reported status as “former”, “current smoker’’ or “never smoker”. Dementia diagnosis was collected by screening at each visit and through continuous automated linkage with digitized medical records and regional registries (**Supplemental Data 2**). Death certificates were obtained via municipal population registries and through general practitioners’ and hospitals’ databases, with complete linkage. This ascertainment method means the Rotterdam Study has functionally no loss to follow-up with respect to dementia diagnosis and death.

Individuals ages 55-70 years who reported smoking (current or former) and who did not have history of dementia at cohort entry were eligible for the current study. To emulate the ideal trial described in Section 3, we contrast former and current smokers. This contrast has some limitations when viewed as an emulation of the ideal trial described in Section 2. For example, there may be unmeasured confounding, selection bias due to misaligning “time zero”[36,37], and measurement error[19]. A thorough consideration of these other issues would be critical for evaluating the effect size of smoking cessation on dementia risk, but go beyond the scope of this exercise. For didactic purposes, we therefore focus our discussion on how the competing event of death affects the interpretation, analytic decisions, and assumptions evoked.

### 4.1. Methods

To estimate the total effect of smoking cessation on dementia risk, we compared a weighted Aalen-Johannsen estimator in current versus former smokers with weights defined as a product of inverse probability of treatment weights[19] to adjust for the following possible confounders: age at study entry, sex, APOE ε4 status, and educational attainment. Briefly, the weight for a current smoker is defined as the inverse of the probability of smoking conditional on confounders, and for a former smoker as the inverse of quitting conditional on covariates. We estimated these probabilities with a logistic regression model for smoking as a function of the above-mentioned covariates. Specific modeling specifications and weights assessment are presented as **Supplemental Data 3**.

To estimate the controlled direct effect, we compared the complement of a weighted Kaplan-Meier survival estimator in smokers versus former smokers with time indexed in years. The weights in this case are time-varying by follow-up year, defined as a product of the time-fixed weights above and a year-specific inverse probability of censoring by death weights. For an individual still alive in year t, the time t censoring weight is the product of the inverse probability of surviving in each year prior to t, conditional on measured shared causes of death and dementia (that is, variables such as C in Figure 1). For an individual who has died by time t, the year t censoring weight is zero. We estimated survival probabilities using a logistic regression model for death as a function of baseline and time-varying covariates. Baseline covariates included smoking status, age at study entry, sex, APOE ε4 status, and educational attainment; time-varying covariates included systolic blood pressure, BMI, and prevalent and incident comorbid heart disease, cancer, stroke, and diabetes.

We also estimated the total effect of smoking on mortality risk applying the Kaplan-Meier estimator with the weights calculated for handling confounding. We therefore are assuming the same set of measured confounders used to estimate the total effect of smoking on dementia risk are sufficient for addressing confounding of the total effect of smoking on mortality risk.

Estimates of the total and controlled direct effect at 20 years of follow-up are presented as risk differences (RD) and risk ratios (RR). All 95% confidence intervals were calculated using percentile-based bootstrapping based on 500 bootstrap samples. All analysis were performed using R, code is available in https://github.com/palolili23/competing_risks_dementia.

#### Standard protocol approvals, registrations, and patient consents

The Rotterdam Study has been approved by the Medical Ethics Committee of the Erasmus MC (registration number MEC 02.1015) and by the Dutch Ministry of Health, Welfare and Sport (Population Screening Act WBO, license number 1071272-159521-PG). The Rotterdam Study has been entered into the Netherlands National Trial Register (NTR; www.trialregister.nl) and into the WHO International Clinical Trials Registry Platform (ICTRP; www.who.int/ictrp/network/primary/en/) under shared catalogue number NTR6831. All participants provided written informed consent to participate in the study and to have their information obtained from treating physicians.

#### Data availability

Rotterdam Study can be obtained via requests directed toward the management team of the Rotterdam Study, which has a protocol for approving data requests. Because of restrictions based on privacy regulations and informed consent of the participants, data cannot be made freely available in a public repository.

### 4.2. Results

Out of 10994 individuals included in the Rotterdam Study, 4179 individuals met eligibility criteria (55-70 years who reported smoking history at baseline and who did not have history of dementia at study entry). The mean age was 62 years and 1870 (44.7%) were women (**Table 2**). In total, 368 (8.8%) developed dementia and 1318 (31.5%) died over 20 years of follow-up. The median time to dementia was 15.5 years and the median time to death was 13.1 years. Overall, from 1572 who were current smokers at baseline, 117 (7.4%) developed dementia and 630 (40.1%) died; of the 2607 former smokers, 251 (9.6%) developed dementia and 688 (26.4%) died.

**Table 2.** Descriptive characteristics of former and current smokers in the Rotterdam Study.

| Characteristics | Former smokers | Current smokers |
| --- | --- | --- |
|  | n = 2607 | n = 1572 |
| Age, mean years (SD) | 62.35 (4.0) | 61.69 (4.0) |
| Women (%) | 1090 (41.8) | 780 (49.6) |
| Education (%) |  |  |
| Primary education | 258 (9.9) | 198 (12.6) |
| Lower or intermediate general education OR lower vocational education | 1080 (41.4) | 693 (44.1) |
| Intermediate vocational education OR higher general education | 862 (33.1) | 483 (30.7) |
| Higher vocational education OR university | 399 (15.3) | 190 (12.1) |
| Unknown | 8 (0.3) | 8 (0.5) |
| APOE-ε4 (%) |  |  |
| Non-carrier | 1747 (67.0) | 1074 (68.3) |
| One allele carrier | 687 (26.4) | 380 (24.2) |
| Two allele carrier | 71 (2.7) | 33 (2.1) |
| Unknown | 102 (3.9) | 85 (5.4) |
| Systolic blood pressure, mean mmHg (SD) | 137.59 (20.8) | 135.22 (21.3) |
| Body mass index, mean (SD) | 26.93 (3.7) | 25.86 (3.8) |
| Prevalent hypertension diagnosis | 1468 (56.3) | 767 (48.8) |
| Prevalent stroke (%) | 52 (2.0) | 23 (1.5) |
| Prevalent heart disease diagnosis (%) | 226 (8.7) | 72 (4.6) |
| Unknown heart disease diagnosis (%) | 42 (1.6) | 28 (1.8) |
| Prevalent diabetes diagnosis (%) | 275 (10.5) | 147 (9.4) |
| Unknown diabetes diagnosis (%) | 389 (14.9) | 364 (23.2) |
| Prevalent cancer diagnosis (%) | 69 (2.6) | 27 (1.7) |

SD: Standard deviation

We estimated a total effect of smoking cessation (compared to continued smoking) on 20-year dementia risk of 2.1 (95%CI: −0.1, 4.2) percentage points (**Table 3; Figure 2**). This slightly harmful effect estimate of quitting smoking (with wide confidence intervals) includes all causal pathways, including that through death. The presence of this pathway is evidenced in the estimated total effect of quitting smoking on 20-year mortality risk: −17.4 (95%CI: −20.5, −14.5) percentage points. Alternatively, we estimated a controlled direct effect of quitting smoking on 20-year dementia risk had death been fully prevented during the study period as −1.9 (−5.1, 1.4) percentage points.

**Table 3.**
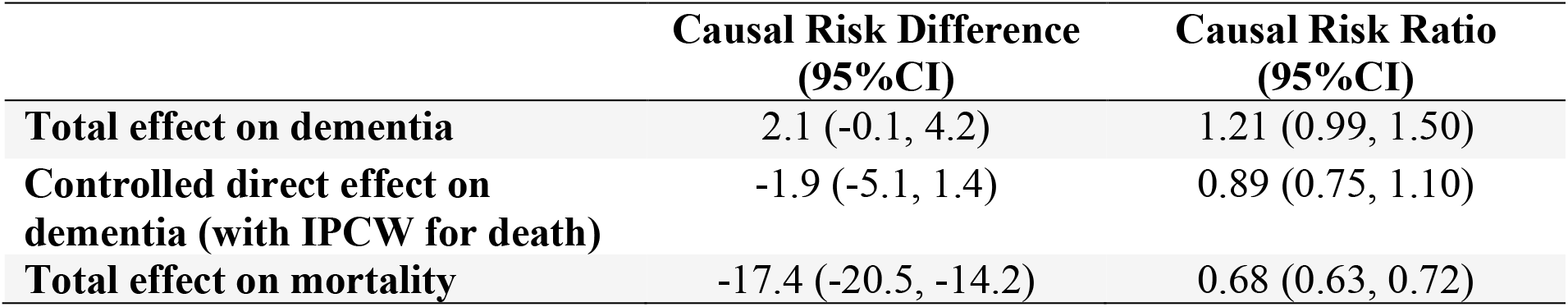
**Total effect and controlled direct effect of smoking cessation (compared to continued smoking) on the risk of dementia, and the total effect on risk of mortality, at 20 years of follow-up**

**Figure 2.**
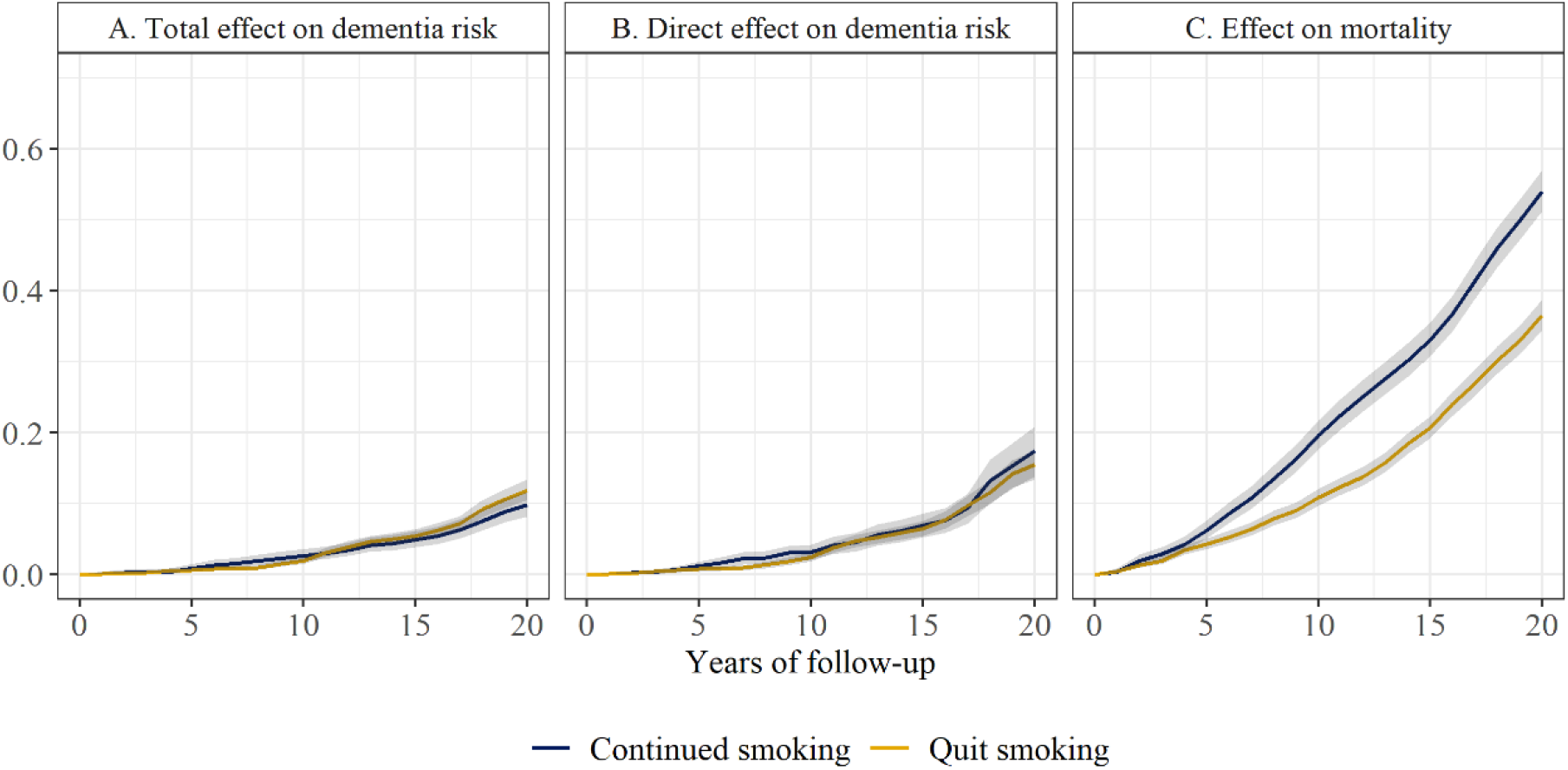
Risk of dementia and death by smoking cessation status over 20 years of follow-up.

## 5. DISCUSSION

In longitudinal (randomized and observational) studies where dementia is the main outcome and deaths occur during follow-up, understanding the different causal questions and making explicit the assumptions required for answering them can lead to better interpretation of results, and a deeper understanding about plausible sources and magnitudes of bias. We considered two causal questions that can be addressed with common statistical methods, beginning with the total effect which captures all causal pathways including those mediated by death. This makes the total effect difficult to interpret as illustrated in our example: the small estimated harmful total effect of smoking cessation on dementia risk necessarily captures some “protection” against dementia by death. *This is not a “bias” but rather a problematic feature of the total effect as the research question*.

The controlled direct effect does not have this problematic feature, and in our example, we estimated that smoking cessation reduced the risk of dementia if death was eliminated. However, residual bias from failing to adjust for a sufficient set of shared causes of death and dementia can remain. In addition, the independent censoring assumption cannot be verified empirically, though bounding can be used to assess extreme scenarios of dependency[3,30,38–40]. Furthermore, the controlled direct effect is not an ideal measure of direct effect because it refers to a fictional scenario where everyone remains alive and therefore it generally will not provide useful information for decision-making.

Since both of these questions can seem unsatisfactory, we note that there are yet further alternative questions that can potentially be posed. For example, the so-called “survivor average treatment effect” quantifies the effect of a treatment on a subgroup of individuals who would not die during the study period under either level of treatment. Although this option has been widely considered in the methodology literature[22,41], the utility of this question is questionable in public health and clinical dementia research as this subgroup is not observable and may not even exist. One can also consider a combined outcome endpoint, such as the effect on dementia *or* death, but this too is unsatisfactory in many cases: in our example, the effect of smoking on risk of death would overwhelm. A novel alternative, the so-called separable effects avoid evoking consideration of implausible scenarios that “eliminate death” or unobservable subpopulations[21]. The separable effects are effects of modified treatments that are assumed to operate like the study treatment but with particular mechanisms removed. These may be a physical decomposition of the exposure assumed to operate on dementia and death through separate pathways or completely different treatments that operate like the study treatment. While identifying these effects similarly rely on strong assumptions, including necessitating measuring a rich set of shared causes of dementia and death, estimates of these effects can be confirmed with future studies on these modified treatments. In this work we focused on the two questions because of their relation to commonly used estimators in dementia research, but we suspect in the future that separable effects will become a more explicit question of interest with the increasing development of useable tools for aiding reasoning.

Too often, we start by defining the statistical method that appears to fit the complexity of data, and we let this decision define the target parameter and thus implicitly determine the research question to be answered. In a setting with competing events, there is no “one size fits all”. Through our discussion and application, we hope that readers will see an opportunity to re-conceptualize how to ask clearer questions in the context of competing events and let the question define the methods that best suit the research aim.

