## Supplemental material for "Choosing questions before methods in dementia research with competing events and causal goals"

**Supplemental Data**

### 1. Systematic Review

#### *a. Searching criteria:*

Search (((((((((("Neurology"[Journal] OR "JAMA"[Journal]) OR "JAMA neurology"[Journal]) OR "Lancet (London, England)"[Journal]) OR "The Lancet. Neurology"[Journal]) OR "Brain : a journal of neurology"[Journal]) OR "Annals of neurology"[Journal]) OR "Alzheimer's & dementia : the journal of the Alzheimer's Association"[Journal]) OR "The New England journal of medicine"[Journal]) OR "BMJ (Clinical research ed.)"[Journal]) AND ((("alzheimer disease"[MeSH Major Topic] OR "dementia"[All fields]) AND (longitudinal[All Fields] OR "cohort studies"[MeSH Terms] OR "cohort"[All Fields])) AND (hazard[All Fields] OR ("risk"[MeSH Terms] OR "risk"[All Fields]))) AND ("2019-01-01"[PDAT] : "2020-01-01"[PDAT])))

#### b. *Flowchart of paper selection for systematic review*

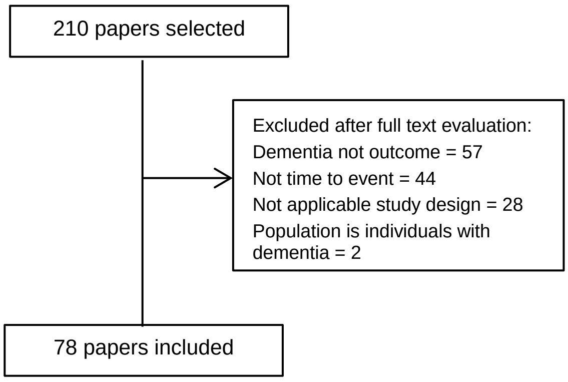

#### *c. Secondary summary measures*

| Values | n |
|---|---|
| Kaplan Meier curves | 18 |
| Rates (cases per person-year) – rate differences | 17 |
| Sub-distribution hazard ratios | 13 |
| Adjusted cumulative incidence/survival curves | 7 |
| C-statistic | 3 |
| Cumulative hazards | 3 |
| Area under the curve graph | 2 |
| Sensitivity and Specificity | 2 |
| Adjusted incidence rates | 1 |
| Age-adjusted risks | 1 |
| Age specific rates considering death | 1 |
| Akaike Information Criterion | 1 |
| Cumulative incidence curves | 1 |
| Life expectancy | 1 |
| Log-hazards by continuous exposure plot | 1 |
| Net Reclassification Index | 1 |
| Observed and predicted Kaplan Meier curves | 1 |
| Population-attributable risk | 1 |
| Smoothed hazard function over time plot | 1 |

### 2. The Rotterdam Study, outcome assessments

*Dementia diagnosis*: Diagnosis was collected by screening during the five study visits, using MMSE and the Geriatric Mental Schedule (GMS) organic level. Screen-positives (MMSE<26 or GMS organic level>0) subsequently underwent an examination and informant interview with the Cambridge Examination for Mental Disorders in the Elderly. A consensus panel led by a consultant neurologist established the final diagnosis according to standard criteria for dementia (DSM-III-R). Additionally, participants were continuously followed for the occurrence of dementia through automated linkage of the study database and digitized medical records from general practitioners and the Regional Institute for Outpatient Mental Health Care. For participants who moved outside the study district or lived in nursing homes, medical records were regularly checked by contacting their treating physicians. Research physicians reviewed all potential dementia cases using hospital discharge letters and information from general practitioners and nursing home physicians[3]. Linkage-based diagnoses were based on data up through December 2015.

*Vital status*: Vital status was obtained on a weekly basis via municipal population registries and through general practitioners' and hospitals' databases, with complete linkage up through December 2015.

**3. Modeling specifications for inverse probability weighting**

| | **Numerator** | **Denominator** |
|---|---|---|
| **Inverse probability of treatment weights**<br><br>$SW^A = \frac{f(A)}{f(A\|A, L)}$ | Pr[A = 1\|1]<br><br>Where:<br>*A* = smoking (0 = current, 1 = former) | Pr[A = 1\|L]<br><br>Where:<br>*A* = smoking (0 = current, 1 = former); *L* = age at study entry with natural cubic splines, sex (women vs. men), education (five categories), apoe4 (four categories), cohort (two categories), and no product terms between covariates |
| **Inverse probability of censoring weights for death**<br><br>$SW^C = \frac{f(C\|A, V)}{f(C\|A, L)}$ | Pr[$D_{k+1}$ = 0\| $D_k$ = 0, A, AY, Y, L]<br><br>Where:<br>*A* = smoking (0 = current, 1 = former); *Y* = year with natural cubic splines; *V* = age at study entry with cubic splines, sex (women vs. men), education (five categories), apoe (four categories), cohort (two categories) and no product terms between covariates | Pr[$D_{k+1}$ = 0\| $D_k$ = 0, A, AY, V, L]<br><br>Where *A* = smoking (0 = current, 1 = former); *Y* = year with natural cubic splines; *V* = age at study entry with cubic splines, sex (women vs. men), education (five categories), apoe (four categories), cohort (two categories); *L* = prevalent diabetes (yes, no), baseline blood pressure with cubic splines, baseline BMI with cubic splines, prevalent hypertension (yes, no), indicator for incident cancer (yes, no), incident heart disease (yes, no), incident diabetes (yes, no) and incident stroke (yes, no) and no product terms between covariates |
